# Effect of COVID-19 infection related experiences on outing behaviors when a state of emergency is declared: a cohort study

**DOI:** 10.1101/2021.08.20.21262364

**Authors:** Takahiro Mori, Tomohisa Nagata, Kazunori Ikegami, Ayako Hino, Seiichiro Tateishi, Mayumi Tsuji, Shinya Matsuda, Yoshihisa Fujino, Koji Mori, the CORoNaWork project

## Abstract

**Background:** Restricting the movement of the public to gathering places and limiting close physical contact are effective measures against COVID-19 infection. In Japan, states of emergency have been declared in specific prefectures to reduce public movement and control COVID-19 transmission. We investigated how COVID-19 infection related experiences including people with a history of infection, people with a history of close contact, and people whose acquaintances have been infected, affected self-restraint related to outing behaviors during the second state of emergency in Japan.

**Methods:** A prospective cohort study was conducted among workers aged 20–65 years using data from an internet survey. The baseline survey was conducted on December 22–25, 2020, and a follow-up survey was on February 18–19, 2021. There were 19,941 participants who completed both surveys and were included in the final analysis. We identified eight outing behaviors: (1) eating out (4 people or fewer); (2) eating out (5 people or more); (3) gathering with friends and colleagues; (4) day trip; (5) overnight trip (excluding visiting home); (6) visiting home; (7) shopping for daily necessities; and (8) shopping for other than daily necessities. We set self-restraint regarding each outing behavior after the second state of emergency was declared in January 2021 as the dependent variable, and COVID-19 infection related experiences as independent variables. Odds ratios were estimated using multilevel logistic regression analyses.

**Results:** Significant differences by COVID-19 infection related experiences were identified: compared to people without COVID-19 related experiences, people with a history of COVID-19 were less likely self-restraint from most outing behaviors. People whose acquaintance had been diagnosed with COVID-19 were significantly more likely to refrain from most outing behaviors. There was no significant difference in any outing behaviors for people with a history of close contact only.

**Conclusions:** To maximize the effect of a state of emergency, health authorities should disseminate information for each person in the target population, taking into account potential differences related to the COVID-19 infection related experiences.

## Introduction

The coronavirus disease 2019 (COVID-19) has been spreading worldwide since 2019. The known routes of COVID-19 infection include droplet infection, aerosol infection, and contact infection, so many infections occur in places where people gather or are in close physical contact [1].

One of effective control measures for COVID-19 infection is to reduce opportunities for people to go to places where people gather or have close physical contact with others [1,2], and the most powerful measure is a restriction of behavior, the so-called lockdown. Lockdowns policies were taken in many countries, although there are differences in methods and degrees of severity. In Japan, the relatively less strict method is to declare a state of emergency and request the citizens to refrain from outing and social gathering. By the end of 2021, a total of four states of emergency have been declared due to the epidemic situation of COVID-19; the first April thru May 2020 against the first wave, the second January thru March 2021against the third wave arrived; the third April thru June 2021 against the fourth wave, and the fourth July thru September 2021 against the fifth wave [3].

The effectiveness of such lockdown policies and states of emergency is likely to be influenced by how much the citizens actually refrain from behavior such as outing and gathering. Some studies have been conducted on sociodemographic factors that influence on outing behavior. Women, older people, highly educated people, and high-income earners are reported to more likely to refrain from outing behavior during a lockdown [4,5,6,7,8,9,10,11,12,13,14]. However, it has been reported that marital status, and whether they have an underlying disease are not associated with outing behaviors during a lockdown [4,5,9,10,11]. In Japan, reports noted that women and young people, people living with family, low-income earners, unemployed people, and those with an underlying disease are more likely to refrain from outing behavior during a state of emergency [15,16], so some factors differ from the reports of other countries during a lockdown.

COVID-19 infection related experiences can be classified into (1) people with a history of infection, (2) people with a history of close contact, and (3) people with acquaintances who have been infected. It was reported people who thought they had had COVID-19 were more likely to think that they had some immunity to the virus and were less likely to adhere to social distancing measures [9]. Conversely, people with a history of close contact or acquaintances may think that they were likely to be infected and further strengthen the measures against infection and refrain from outing behaviors, but these effects have not yet been investigated to our knowledge.

As the pandemic continues, more and more citizens in every country are becoming infected, and the proportion of those with opportunities for intense contact or experience with an infected acquaintance is increasing. Therefore, understanding the association between COVID-19 infection related experiences and self-restraint in behaviors during a state of emergency may have important implications for policy decisions during COVID-19 and other pandemic outbreaks. We therefore investigated how each type of COVID-19 infection related experience affected self-restraint in outing behaviors using the data in January 2021 when the second state of emergency was declared in Japan.

## Methods

### Study design and participants

This prospective cohort study was undertaken by a research group from the University of Occupational and Environmental Health, Japan, called the Collaborative Online Research on Novel-coronavirus and Work study (CORoNaWork study). This survey was conducted as a self-administrated questionnaire by the internet survey company Cross Marketing Inc. (Tokyo, Japan). The baseline survey was conducted on December 22–25, 2020, and the follow-up survey was on February 18–19, 2021; both periods were during the third wave of the pandemic in Japan. Details of the study protocol have been previously reported [17]. Participants (n = 33,087) were aged 20–65 years and employed at the time of the baseline survey. Respondents to the CORoNaWork study were sampled taking into account region, occupation, and sex. After excluding 6,051 initial subjects who provided invalid responses, we ultimately included 27,036 in the database. Invalid responses were determined as follows: response time <6 minutes, body weight <30 kg, height <140 cm, inconsistent answers to similar questions, and wrong answers to a question used solely to identify unreliable responses. These subjects were given a follow-up survey, and 19,941 people responded (74% follow-up rate). The flow diagram is shown in Figure 1.

**Figure 1.**
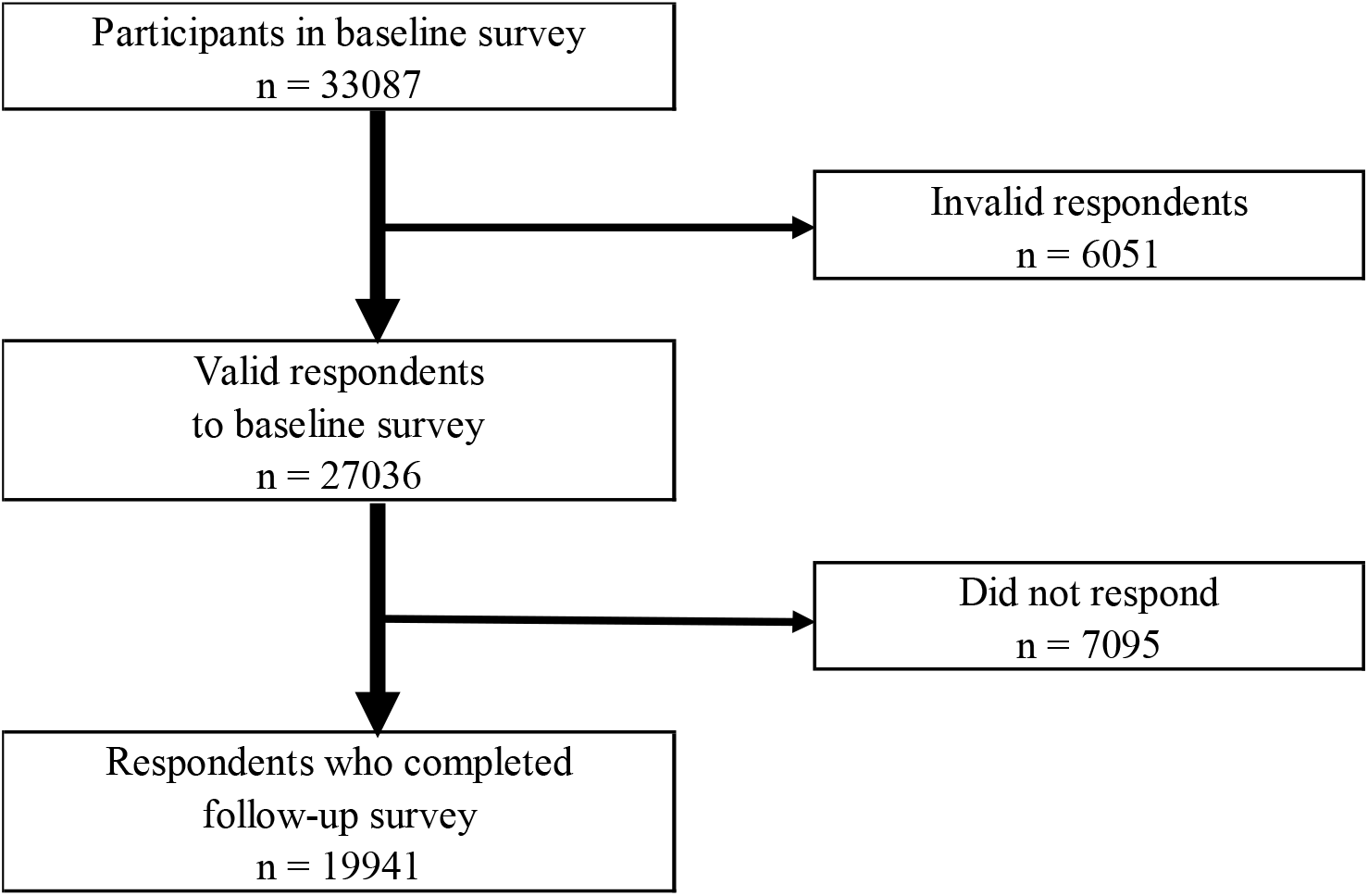
Flow chart of the study participants.

The present study was approved by the Ethics Committee of the University of Occupational and Environmental Health, Japan (Approval numbers: R2-079 and R3-006). Informed consent was obtained in the form of the website from all participants.

### Assessment of outing behaviors

We identified eight outing behaviors which the Japanese government requested for self-restraint [18]: (1) eating out (4 people or fewer); (2) eating out (5 people or more); (3) gathering with friends and colleagues; (4) day trip; (5) overnight trip (excluding visiting home); (6) visiting home; (7) shopping for daily necessities; and (8) shopping for other than daily necessities. We divided eating out into four people or fewer and five or more to identify any difference based on the number of people, particularly because the Japanese government has emphasized that drinking or dining with five or more people carries a particularly high risk of infection [18]. For each of the eight behaviors, we asked participants in the follow-up survey, “Has your self-restraint changed in response to the second state of emergency in January 2021?” Respondents chose one of the following five options: very increased, a little increased, no change, a little decreased, or very decreased. We created a binary variable by defining “a little decreased” or “very decreased” as having self-restraint behavior, and the others as not having self-restraint behavior.

### Assessment of the COVID-19 related experiences

In the baseline survey, we asked participants three questions about their COVID-19 infection related experiences: “Have you ever been infected with COVID-19?”, “Have you ever been in close contact with someone with COVID-19?”, and “Do you have an acquaintance who has been infected with COVID-19?” Respondents answered each question with “Yes” or “No”, and were classified into the four following types: people with a history of COVID-19; people without history of COVID-19 but with a history of close contact with cases of confirmed COVID-19 (hereinafter referred to as people with a history of close contact); people without a history of COVID-19 or close contact with cases of confirmed COVID-19 but who had an acquaintance who had been diagnosed with COVID-19 (hereinafter referred to as people whose acquaintance had been diagnosed); and people without history of COVID-19 or close contact with cases of confirmed COVID-19, and who did not have an acquaintance diagnosed with COVID-19 (hereinafter referred to as people without history of COVID-19 or close contact).

### Assessment of covariates

Covariates included demographics, socioeconomic factors, job type, underlying disease, and prefectures with and without the second state of emergency. Age was expressed as a continuous variable. Education was classified into three categories: junior high or high school, vocational school or college, and university or graduate school. Marital status was classified into three categories: married, divorced or widowed, and never married. Equivalent income was classified into four categories: <2.50 million Japanese yen (JPY); 2.50–3.74 million JPY; 3.75–5.24 million JPY; and ≥5.25 million JPY (1 USD was equal to 106.78 JPY, using 2020 conversion rates) [19]. Job type was classified into three categories: mainly desk work, jobs mainly involving interpersonal communication, and mainly physical work. Regarding underlying disease, we asked the question, “Do you have any disease that requires regular visits to the hospital or treatment?” Participants selected one of the following: “I do not have such a disease,” “I have such a disease,” or “I have such a disease, but refrained from going to the hospital following the second state of emergency.” We rated the participants who answered, “I do not have such a disease” as “No” and the remaining two answers as “Yes.” There were 11 prefectures that had the second state of emergency, and 36 prefectures without this declaration.

### Statistical analyses

Multilevel logistic regression analyses were used to examine the association between COVID-19 infection related experience and self-restraint from outing behaviors after the second state of emergency. An analysis was performed on each of the eight outing behaviors. We estimated age-sex adjusted odds ratios (ORs) and multivariate adjusted ORs for each outing behaviors using multilevel logistic regression analyses nested in the prefecture of residence to take account of regional differences in the infection status of COVID-19. The model included age, sex, education, marital status, equivalent income, job type, underlying disease, and prefectures with and without the second state of emergency. A p-value of less than 0.05 was considered statistically significant. All analyses were conducted using Stata Statistical Software (Release 16; StataCorp LLC, College Station, TX, USA).

## Results

Table 1 shows the participant characteristics for each category of COVID-19 infection related experience. There were 18,208 people without history of COVID-19 or close contact, 154 people with a history of COVID-19, 141 people with a history of close contact, and 1438 people whose acquaintance had been diagnosed. The mean age was youngest for people with a history of COVID-19. People without history of COVID-19 or close contact had a lower rate of being married than did other groups. In prefectures with the emergency declaration, there were fewer people without history of COVID-19 or close contact and more people with a history of COVID-19, with a history of close contact, and whose acquaintance was diagnosed than in prefectures without the declaration.

**Table 1.** Characteristics of participants by categories of COVID-19 infection related experience

|  | People without<br>history of<br>COVID-19 or<br>close contact*<br>n=18208 | People with a<br>history of<br>COVID-19<br>n=154 | People with a<br>history of<br>close contact†<br>n=141 | People whose<br>acquaintance<br>had been<br>diagnosed‡<br>n=1438 |
| --- | --- | --- | --- | --- |
| Age, mean (SD) | 48.0 (10.1) | 43.8 (11.5) | 45.7 (10.6) | 47.9 (10.2) |
| Sex, men | 10195 (56.0%) | 85 (55.2%) | 87 (61.7%) | 803 (55.8%) |
| Education |  |  |  |  |
| Junior high or high school | 4951 (27.2%) | 37 (24.0%) | 23 (16.3%) | 349 (24.3%) |
| Vocational school or college | 4221 (23.2%) | 35 (22.7%) | 37 (26.2%) | 292 (20.3%) |
| University or graduate school | 9036 (49.6%) | 82 (53.2%) | 81 (57.4%) | 797 (55.4%) |
| Marital status |  |  |  |  |
| Married | 10134 (55.7%) | 96 (62.3%) | 85 (60.3%) | 870 (60.5%) |
| Divorced or bereaved | 1848 (10.1%) | 14 (9.1%) | 10 (7.1%) | 164 (11.4%) |
| Never married | 6226 (34.2%) | 44 (28.6%) | 46 (32.6%) | 404 (28.1%) |
| Equivalent income |  |  |  |  |
| <2.50 million JPY | 3976 (21.8%) | 33 (21.4%) | 17 (12.1%) | 207 (14.4%) |
| 2.50-3.74 million JPY | 4588 (25.2%) | 35 (22.7%) | 31 (22.0%) | 344 (23.9%) |
| 3.75-5.24 million JPY | 4822 (26.5%) | 37 (24.0%) | 38 (27.0%) | 384 (26.7%) |
| ≥5.25 million JPY | 4822 (26.5%) | 49 (31.8%) | 55 (39.0%) | 503 (35.0%) |
| Job type |  |  |  |  |
| Mainly desk work | 9381 (51.5%) | 72 (46.8%) | 61 (43.3%) | 754 (52.4%) |
| Jobs mainly involving<br>interpersonal communication | 4425 (24.3%) | 46 (29.9%) | 44 (31.2%) | 422 (29.3%) |
| Mainly physical work | 4402 (24.2%) | 36 (23.4%) | 36 (25.5%) | 262 (18.2%) |
| Underlying disease, Yes | 6346 (34.9%) | 73 (47.4%) | 60 (42.6%) | 559 (38.9%) |
| Prefectures with the second state of<br>emergency, Yes | 7746 (42.5%) | 81 (52.6%) | 92 (65.2%) | 783 (54.5%) |

\* People without history of COVID-19 or close contact with cases of confirmed COVID-19, and whose acquaintance was not diagnosed with COVID-19
† People with a history of close contact with cases of confirmed COVID-19 and without history of COVID-19
‡ People without a history of COVID-19 or close contact with confirmed cases of COVID-19 whose acquaintance had been diagnosed with COVID-19
SD, standard deviation; JPY, Japanese yen

Table 2 shows the association between COVID-19 infection related experiences and self-restraint from outing behaviors during the second state of emergency. In multivariate analysis adjusted for age, sex, education, marital status, equivalent income, job type, underlying disease, and prefectures with and without the second state of emergency, people with a history of COVID-19 were less likely self-restraint from eating out (4 people or less) (OR 0.47), eating out (5 people or more) (OR 0.48), gathering with friends and colleagues (OR 0.39), day trip (OR 0.49), overnight trip (OR 0.48), visiting home (OR 0.70) and shopping other than daily necessities (OR 0.65) than people without history of COVID-19 or close contact, but there was no significant difference in shopping for daily necessities. Conversely, people whose acquaintance had been diagnosed was significantly more likely self-restraint from eating out (4 people or fewer) (OR 1.37), eating out (5 people or more) (OR 1.45), gathering with friends and colleagues (OR 1.52), day trip (OR 1.34), overnight trip (OR 1.35), visiting home (OR 1.21), and shopping other than daily necessities (OR 1.29) than people without history of COVID-19 and close contact, but there was no significant difference for shopping for daily necessities. People with a history of close contact showed no significant difference in any of the outing behaviors compared with people without history of COVID-19 and close contact.

**Table 2.** Association between COVID-19 infection related experience and self-restraint from outing behaviors during the second state of emergency

|  | Self-restraint from<br>outing behaviors |  | Age-sex adjusted |  |  | Multivariate adjusted* |  |  |
| --- | --- | --- | --- | --- | --- | --- | --- | --- |
|  | n | % | OR | 95% CI | P | OR | 95% CI | P |
| (1) Eating out (4 people or fewer) |  |  |  |  |  |  |  |  |
| People without history of COVID-19 or close contact† | 9921 | 54 | reference |  |  | reference |  |  |
| People with a history of COVID-19 | 60 | 39 | 0.50 | 0.36-0.70 | <0.001 | 0.47 | 0.34-0.65 | <0.001 |
| People with a history of close contact‡ | 86 | 61 | 1.25 | 0.89-1.76 | 0.207 | 1.15 | 0.81-1.62 | 0.429 |
| People whose acquaintance had been diagnosed§ | 924 | 64 | 1.45 | 1.30-1.63 | <0.001 | 1.37 | 1.22-1.53 | <0.001 |
| (2) eating out (5 people or more) |  |  |  |  |  |  |  |  |
| People without history of COVID-19 or close contact† | 10516 | 58 | reference |  |  | reference |  |  |
| People with a history of COVID-19 | 65 | 42 | 0.53 | 0.38-0.73 | <0.001 | 0.48 | 0.34-0.66 | <0.001 |
| People with a history of close contact‡ | 90 | 64 | 1.26 | 0.89-1.79 | 0.184 | 1.14 | 0.80-1.62 | 0.467 |
| People whose acquaintance had been diagnosed§ | 986 | 69 | 1.56 | 1.39-1.76 | <0.001 | 1.45 | 1.29-1.63 | <0.001 |
| (3) gathering with friends and colleagues |  |  |  |  |  |  |  |  |
| People without history of COVID-19 or close contact† | 11904 | 65 | reference |  |  | reference |  |  |
| People with a history of COVID-19 | 69 | 45 | 0.44 | 0.32-0.60 | <0.001 | 0.39 | 0.28-0.54 | <0.001 |
| People with a history of close contact‡ | 92 | 65 | 1.01 | 0.71-1.44 | 0.938 | 0.90 | 0.63-1.28 | 0.560 |
| People whose acquaintance had been diagnosed§ | 1090 | 76 | 1.65 | 1.45-1.87 | <0.001 | 1.52 | 1.34-1.72 | <0.001 |
| (4) day trip |  |  |  |  |  |  |  |  |
| People without history of COVID-19 or close contact† | 10740 | 59 | reference |  |  | reference |  |  |
| People with a history of COVID-19 | 68 | 44 | 0.54 | 0.39-0.75 | <0.001 | 0.49 | 0.35-0.68 | <0.001 |
| People with a history of close contact‡ | 86 | 61 | 1.09 | 0.78-1.54 | 0.617 | 0.98 | 0.69-1.38 | 0.890 |
| People whose acquaintance had been diagnosed§ | 972 | 68 | 1.44 | 1.28-1.62 | <0.001 | 1.34 | 1.19-1.50 | <0.001 |

\* Adjusted for age, sex, education, marital status, equivalent income, job type, underlying disease, and prefectures with and without the second state of emergency
† People without history of COVID-19 or close contact with cases of confirmed COVID-19, and whose acquaintance was not diagnosed with COVID-19
‡ People with a history of close contact with cases of confirmed COVID-19 and without history of COVID-19
§ People without a history of COVID-19 or close contact with confirmed cases of COVID-19 whose acquaintance had been diagnosed with COVID-19

Table 2. (continued)
|  | Self-restraint from<br>outing behaviors |  | Age-sex adjusted |  |  | Multivariate adjusted* |  |  |
| --- | --- | --- | --- | --- | --- | --- | --- | --- |
|  | n | % | OR | 95% CI | P | OR | 95% CI | P |
| (5) overnight trip (excluding visiting home) |  |  |  |  |  |  |  |  |
| People without history of COVID-19 or close contact† | 10983 | 60 | reference |  |  | reference |  |  |
| People with a history of COVID-19 | 69 | 45 | 0.53 | 0.39-0.74 | <0.001 | 0.48 | 0.35-0.66 | <0.001 |
| People with a history of close contact‡ | 90 | 64 | 1.17 | 0.83-1.65 | 0.375 | 1.03 | 0.73-1.47 | 0.855 |
| People whose acquaintance had been diagnosed§ | 994 | 69 | 1.46 | 1.30-1.64 | <0.001 | 1.35 | 1.20-1.52 | <0.001 |
| (6) visiting home |  |  |  |  |  |  |  |  |
| People without history of COVID-19 or close contact† | 8539 | 47 | reference |  |  | reference |  |  |
| People with a history of COVID-19 | 65 | 42 | 0.78 | 0.56-1.08 | 0.131 | 0.70 | 0.50-0.97 | 0.030 |
| People with a history of close contact‡ | 75 | 53 | 1.19 | 0.85-1.66 | 0.316 | 1.07 | 0.76-1.51 | 0.691 |
| People whose acquaintance had been diagnosed§ | 785 | 55 | 1.30 | 1.17-1.45 | <0.001 | 1.21 | 1.08-1.35 | 0.001 |
| (7) shopping for daily necessities |  |  |  |  |  |  |  |  |
| People without history of COVID-19 or close contact† | 4980 | 27 | reference |  |  | reference |  |  |
| People with a history of COVID-19 | 53 | 34 | 1.33 | 0.95-1.87 | 0.096 | 1.28 | 0.91-1.80 | 0.149 |
| People with a history of close contact‡ | 45 | 32 | 1.24 | 0.87-1.78 | 0.235 | 1.20 | 0.84-1.72 | 0.313 |
| People whose acquaintance had been diagnosed§ | 437 | 29 | 1.09 | 0.97-1.23 | 0.160 | 1.06 | 0.94-1.20 | 0.303 |
| (8) shopping for other than daily necessities |  |  |  |  |  |  |  |  |
| People without history of COVID-19 or close contact† | 7973 | 44 | reference |  |  | reference |  |  |
| People with a history of COVID-19 | 55 | 36 | 0.69 | 0.49-0.97 | 0.031 | 0.65 | 0.46-0.91 | 0.012 |
| People with a history of close contact‡ | 61 | 43 | 0.99 | 0.70-1.38 | 0.931 | 0.92 | 0.65-1.29 | 0.613 |
| People whose acquaintance had been diagnosed§ | 741 | 52 | 1.35 | 1.21-1.50 | <0.001 | 1.29 | 1.15-1.44 | <0.001 |

\* Adjusted for age, sex, education, marital status, equivalent income, job type, underlying disease, and prefectures with and without the second state of emergency
† People without history of COVID-19 or close contact with cases of confirmed COVID-19, and whose acquaintance was not diagnosed with COVID-19
‡ People with a history of close contact with cases of confirmed COVID-19 and without history of COVID-19
§ People without a history of COVID-19 or close contact with confirmed cases of COVID-19 whose acquaintance had been diagnosed with COVID-19

## Discussion

We examined the association of COVID-19 infection related experiences with self-restraint from outing behaviors during the second state of emergency. People with a history of COVID-19 reported significantly lower self-restraint from outing behaviors than did people without a history of COVID-19 or close contact, except regarding shopping for daily necessities. People whose acquaintance had been diagnosed were significantly more likely to refrain from outing behaviors except for shopping for daily necessities. The results of people with a history of COVID-19 and people whose acquaintance had been diagnosed were similar to what we had expected. It has been shown that risk perception for infection is involved in infection prevention behavior [20,21,22].

Risk perception for infection includes perceptions of the possibility and severity of infection [23]. The reason why people with a history of COVID-19 were less likely to refrain from outing behaviors may result from their lower risk perception; they were less likely to be infected with COVID-19 again and less likely to become severe. The association between risk perception and preventive behavior was also reported in various countries during the 2009 H1N1 influenza epidemic [24,25,26,27,28,29,30]. It is consistent with the pervious study observed that people who believed they had had COVID-19 were more likely to report leaving home at early stage of COVID-19 in UK [9]. At the time of this survey, the number of infected people in Japan was less than 0.5% of the total population [18]. This infection rate was so low that infected people might have thought that they were unlucky even once infected, and infected persons often acquire neutralizing antibodies, so they might also have thought that re-infection was unlikely to occur [31,32]. In addition, because past experiences have been reported to be less associated with concerns about voluntary risk behavior [33], a COVID-19 diagnosis might not lead to refraining from outing behaviors that may have caused them to become infected in the past or increase their risk of infection. As for the perception of severity, if their infection was relatively mild or asymptomatic, they may have had perceived the severity of COVID-19 infection to be lesser. Accordingly, even when a state of emergency was declared, there was no change in self-restraint from outing behaviors.

Conversely, people whose acquaintance had been diagnosed were considered to have increased risk perception. A survey of Japanese people showed that the risk to oneself is underestimated than the risk to society when comparing the degree to which one feels dangerous to oneself and the degree to one feels dangerous to society for the same infectious disease [34]. However, if a close acquaintance was infected or even became seriously ill, it is probable that the risk perception to oneself increased, leading to refraining from outing behavior.

For people with a history of close contact, the results were different from our assumptions, and no significant difference was observed in any of outing behaviors. The reasons for this result are not clear from this survey, but it is possible that there were both those who refrained from outing behaviors and those who did not. In Japan, epidemiological surveys are conducted by health centers on each infected person, and close contacts are identified. After certification, close contacts undergo a real-time reverse transcription–polymerase chain reaction (rt-PCR) test to check for infection and are quarantined at home for 14 days at the time of our survey, even if negative [35]. As mentioned above, people with a history of close contact felt close to the infection, so it is probable that they may have adopted the same behaviors as people whose acquaintance had been diagnosed. Conversely, it seems that there were a number of people who did not refrain from outing behavior. Some people may think that they have already been infected because of the possibility of false negatives even if the rt-PCR test is negative, or from the experience of being isolated at home for 14 days. In addition, many people are recognized as close contacts as a result of their family members living together being infected [36,37], and often make decisions about outing behaviors with their family members. These people therefore may have not refrained from outing behaviors like people with a history of COVID-19.

Regarding the specific aspects of outing behaviors, for shopping for daily necessities, neither people with a history of COVID-19 nor people whose acquaintance had been diagnosed showed a significant difference compared with people without history of COVID-19 and close contact. This may be because shopping for daily necessities is a daily activity necessary for daily life, unlike other outing behaviors. Regarding eating out, similar results were seen in eating out with four people or fewer and with five or more, suggesting that they have a similar perception of the risk of infection.

Our results suggested that it is necessary to pay attention not only to sociodemographic factors that have already been investigated [4,5,6,7,8,9,10,11,12,13,14,15,16], but also to the COVID-19 infection related experiences. In particular, our finding that people whose acquaintance had been diagnosed were more likely to refrain from outing behaviors suggested that in order to encourage people without COVID-19-related experiences to properly refrain from outing behaviors, officials need to make these people feel more familiar to the risk of infection. Since it has been shown that the information effect is large [38], it is necessary to disseminate information that makes the infection more familiar according to the actual situation, and also that refraining from outing behaviors is beneficial to society as a whole, including the national health care system. In addition, individual risk preference is known to be influenced by the decision-making of surrounding individuals [39], and it will be easier to make the choice to go out if many people go out when self-restraint is requested. It is therefore necessary to provide appropriate information to people with a history of COVID-19, such as the possibility of re-infection and the effects of new variants of SARS-CoV-2 [40].

There are several limitations to this study. First, we conducted an internet survey, which includes the possibility of selection bias. However, sampling was balanced by sex, occupation, and area of residence at the start of the study to reduce the potential for bias. Second, the follow-up survey was conducted two months after the baseline survey. We classified the COVID-19 infection related experience using data from the baseline survey, so there may have been new people infected in the intervening two months. However, the ratio in each category implies that the number of newly applicable people was very small, and unlikely to significantly affect the results. Third, the outcome of interest in this survey was a decrease in self-restraint, the specific degree of self-restraint was not investigated. In addition, those who were not originally outing at all before the second state of emergency was declared were included in the group who did not respond to self-restraint. However, the movement of people during the non-declaration period does not appear to have been significantly suppressed compared with before the COVID-19 epidemic [41]. This implies that there would have been few people in this category who did not go out at all during the non-declaration period.

## Conclusion

Our results show that the level of self-restraint from outing behaviors due to a state of emergency differs depending on a subject’s experiences related to COVID-19 infection. When declaring a state of emergency, in order to maximize the effect of the declaration, officials should disseminate information in a way that makes the infection feel more familiar.

## Data Availability

Data not available due to ethical restrictions

## Data Availability

Data not available due to ethical restrictions

## Abbreviations

COVID-19: Coronavirus disease 2019
CORoNaWork: Collaborative Online Research on the Novel-coronavirus and Work
JPY: Japanese yen
OR: Odds ratio

## Declarations

### Competing interests

The authors declare that they have no competing interests.

### Funding

This study was supported and partly funded by the research grant from the University of Occupational and Environmental Health, Japan (no grant number); Japanese Ministry of Health, Labour and Welfare (H30-josei-ippan-002, H30-roudou-ippan-007, 19JA1004, 20JA1006, 210301-1, and 20HB1004); Anshin Zaidan (no grant number), the Collabo-Health Study Group (no grant number), and Hitachi Systems, Ltd. (no grant number) and scholarship donations from Chugai Pharmaceutical Co., Ltd. (no grant number)

### Authors’ contributions

TM; data analysis and writing the manuscript, TN; creating the questionnaire, review of manuscript, and advice on interpretation, KI, AH, ST, MT, and SM; review of manuscripts, and advice on interpretation, YF; overall survey planning, creating the questionnaire, review of manuscripts, and advice on interpretation, KM; data analysis, drafting the manuscript, review of manuscripts, and advice on interpretation

## Acknowledgments

The current members of the CORoNaWork Project, in alphabetical order, are as follows: Dr. Yoshihisa Fujino (present chairperson of the study group), Dr. Akira Ogami, Dr. Arisa Harada, Dr. Ayako Hino, Dr. Hajime Ando, Dr. Hisashi Eguchi, Dr. Kazunori Ikegami, Dr. Kei Tokutsu, Dr. Keiji Muramatsu, Dr. Koji Mori, Dr. Kosuke Mafune, Dr. Kyoko Kitagawa, Dr. Masako Nagata, Dr. Mayumi Tsuji, Ms. Ning Liu, Dr. Rie Tanaka, Dr. Ryutaro Matsugaki, Dr. Seiichiro Tateishi, Dr. Shinya Matsuda, Dr. Tomohiro Ishimaru, and Dr. Tomohisa Nagata. All members are affiliated with the University of Occupational and Environmental Health, Japan.

## Notes

### Competing Interest Statement

The authors have declared no competing interest.

### Author Declarations

The present study was approved by the Ethics Committee of the University of Occupational and Environmental Health, Japan (Approval numbers: R2-079 and R3-006).

## References

1. Honein MA, Christie A, Rose DA, Brooks JT, Meaney-Delman D, Cohn A, et al. Summary of Guidance for Public Health Strategies to Address High Levels of Community Transmission of SARS-CoV-2 and Related Deaths, December 2020. MMWR Morb Mortal Wkly Rep 2020;69:1860–7. doi.org/10.15585/mmwr.mm6949e2.

2. Would Health Organization. Transmission of SARS-CoV-2: implications for infection prevention precautions. https://www.who.int/news-room/commentaries/detail/transmission-of-sars-cov-2-implications-for-infection-prevention-precautions. Accessed 29 April 2022.

3. Prime Minister of Japan and His Cabinet. Ongoing Topics. https://japan.kantei.go.jp/ongoingtopics/index.html. Accessed 29 April 2022.

4. Coroiu A, Moran C, Campbell T, Geller AC. Barriers and facilitators of adherence to social distancing recommendations during COVID-19 among a large international sample of adults. PLoS One. 2020;15:e0239795.

5. Gouin JP, MacNeil S, Switzer A, Carrese-Chacra E, Durif F, Knäuper B. Socio-demographic, social, cognitive, and emotional correlates of adherence to physical distancing during the COVID-19 pandemic: a cross-sectional study. Can J Public Health. 2021;112:17–28.

6. Gualda E, Krouwel A, Palacios-Gálvez M, Morales-Marente E, Rodríguez-Pascual I, García-Navarro EB. Social Distancing and COVID-19: Factors Associated With Compliance With Social Distancing Norms in Spain. Front Psychol. 2021;12:727225.

7. Park CL, Russell BS, Fendrich M, Finkelstein-Fox L, Hutchison M, Becker J. Americans’ COVID-19 Stress, Coping, and Adherence to CDC Guidelines. J Gen Intern Med. 2020;35:2296–2303.

8. Clark C, Davila A, Regis M, Kraus S. Predictors of COVID-19 voluntary compliance behaviors: An international investigation. Glob Transit. 2020;2:76–82.

9. Smith LE, Mottershaw AL, Egan M, Waller J, Marteau TM, Rubin GJ. The impact of believing you have had COVID-19 on self-reported behaviour: Cross-sectional survey. PLoS One. 2020;15:e0240399.

10. Al Zabadi H, Yaseen N, Alhroub T, Haj-Yahya M. Assessment of Quarantine Understanding and Adherence to Lockdown Measures During the COVID-19 Pandemic in Palestine: Community Experience and Evidence for Action. Front Public Health. 2021;9:570242.

11. Papageorge NW, Zahn MV, Belot M, van den Broek-Altenburg E, Choi S, Jamison JC, et al. Socio-demographic factors associated with self-protecting behavior during the Covid-19 pandemic. J Popul Econ. 2021:1–48.

12. Turk E, Čelik T, Smrdu M, Šet J, Kuder A, Gregorič M, et al. Adherence to COVID-19 mitigation measures: The role of sociodemographic and personality factors. Curr Psychol. 2021:1–17.

13. Iio K, Guo X, Kong X, Rees K, Bruce Wang X. COVID-19 and social distancing: Disparities in mobility adaptation between income groups. Transp Res Interdiscip Perspect. 2021;10:100333.

14. Pullano G, Valdano E, Scarpa N, Rubrichi S, Colizza V. Evaluating the effect of demographic factors, socioeconomic factors, and risk aversion on mobility during the COVID-19 epidemic in France under lockdown: a population-based study. Lancet Digit Health. 2020;2:e638–e649.

15. Hanibuchi T, Yabe N, Nakaya T. Who is staying home and who is not? Demographic, socioeconomic, and geographic differences in time spent outside the home during the COVID-19 outbreak in Japan. Prev Med Reports. 2021;21:101306.

16. Watanabe T, Yabu T. Japan’s voluntary lockdown: further evidence based on age-specific mobile location data. Japanese Econ Rev. 2021:1–38.

17. Fujino Y, Ishimaru T, Eguchi H, Tsuji M, Tateishi S, Ogami A, et al. Protocol for a nationwide Internet-based health survey in workers during the COVID-19 pandemic in 2020. J UOEH. 2021;43:217–225.

18. Cabinet Secretariat. COVID-19 Information and Resources. https://corona.go.jp/en/dashboard/. Accessed 29 April 2022.

19. OECD. Exchange rates (indicator). https://doi.org/10.1787/037ed317-en. Accessed 29 April 2022.

20. Sutton SR. Social-psychological approaches to understanding addictive behaviours: attitude-behaviour and decision-making models. Br J Addict 1987;82:355–70.

21. Weinstein ND. Testing four competing theories of health-protective behavior. Health Psychol 1993;12:324–33.

22. Brewer NT, Weinstein ND, Cuite CL, Herrington JE. Risk Perceptions and Their Relation to Risk Behavior. Ann Behav Med 2004;27:125–30.

23. Brewer NT, Chapman GB, Gibbons FX, Gerrard M, McCaul KD, Weinstein ND. Meta-analysis of the relationship between risk perception and health behavior: The example of vaccination. Heal Psychol 2007;26:136–45.

24. Jones JH, Salathe M. Early assessment of anxiety and behavioral response to novel swine-origin influenza A(H1N1). PLoS One 2009;4:e8032.

25. Chor JSY, Ngai KLK, Goggins WB, Wong MCS, Wong SYS, Lee N, et al. Willingness of Hong Kong healthcare workers to accept pre-pandemic influenza vaccination at different WHO alert levels: Two questionnaire surveys. BMJ 2009;339:b3391.

26. Rubin GJ, Amlôt R, Page L, Wessely S. Public perceptions, anxiety, and behaviour change in relation to the swine flu outbreak: Cross sectional telephone survey. BMJ 2009;339:b2651.

27. Seale H, Heywood AE, McLaws ML, Ward KF, Lowbridge CP, Van D, et al. Why do I need it? I am not at risk! Public perceptions towards the pandemic (H1N1) 2009 vaccine. BMC Infect Dis 2010;10.

28. Lau JTF, Griffiths S, Choi KC, Tsui HY. Avoidance behaviors and negative psychological responses in the general population in the initial stage of the H1N1 pandemic in Hong Kong. BMC Infect Dis 2010;10.

29. Lau JTF, Griffiths S, Choi K chow, Lin C. Prevalence of preventive behaviors and associated factors during early phase of the H1N1 influenza epidemic. Am J Infect Control 2010;38:374–80.

30. Setbon M, Raude J. Factors in vaccination intention against the pandemic influenza A/H1N1. Eur J Public Health 2010;20:490–4.

31. Hansen CH, Michlmayr D, Gubbels SM, Mølbak K, Ethelberg S. Assessment of protection against reinfection with SARS-CoV-2 among 4 million PCR-tested individuals in Denmark in 2020: a population-level observational study. Lancet 2021;397:1204–12.

32. Yamayoshi S, Yasuhara A, Ito M, Akasaka O, Nakamura M, Nakachi I, et al. Antibody titers against SARS-CoV-2 decline, but do not disappear for several months. EClinicalMedicine 2021;32:100734.

33. Rubin GJ, Amlôt R, Page L, Wessely S. Public perceptions, anxiety, and behaviour change in relation to the swine flu outbreak: Cross sectional telephone survey. BMJ 2009;339:b2651.

34. Inamasu T, Horiguchi I, Marui E. Vulnerability and risk perception to infectious diseases of Japanese [In Japanese]. Journal of health and welfare statistics 2013;60:40–44.

35. National institute of infectious diseases. Guidelines for conducting an active epidemiological survey on patients with COVID-19 infection, November 29, 2021 [In Japanese]. https://www.niid.go.jp/niid/images/cfeir/covid19/COVID19-02-211129.pdf. Accessed 29 April 2022.

36. Koh WC, Naing L, Chaw L, Rosledzana MA, Alikhan MF, Jamaludin SA, et al. What do we know about SARS-CoV-2 transmission? A systematic review and meta-analysis of the secondary attack rate and associated risk factors. PLoS One 2020;15:e0240205.

37. Miyahara R, Tsuchiya N, Yasuda I, Ko YK, Furuse Y, Sando E, et al. Familial Clusters of Coronavirus. Emerg Infect Dis 2021;27:915–918.

38. Watanabe T, Yabu T. Japan’ s Voluntary Lockdown 2020. PLOS ONE 2021;16:e0252468.

39. Chung D, Christopoulos GI, King-Casas B, Ball SB, Chiu PH. Social signals of safety and risk confer utility and have asymmetric effects on observers’ choices. Nat Neurosci 2015;18:912–6.

40. Centers for Disease Control and Prevention. SARS-CoV-2 Variant Classifications and Definitions. https://www.cdc.gov/coronavirus/2019-ncov/variants/variant-info.html. Accessed 29 April 2022.

41. Cabinet Secretariat. COVID-19 Information and Resources: The amount of people flowing between prefectures and their changes [In Japanese]. https://corona.go.jp/dashboard/pdf/flow_20210712.pdf. Accessed 29 April 2022.

